# Estimating the contribution of subclinical tuberculosis disease to transmission – an individual patient data analysis from prevalence surveys

**DOI:** 10.1101/2022.06.09.22276188

**Authors:** Jon C. Emery, Peter J. Dodd, Sayera Banu, Beatrice Frascella, Frances L. Garden, Katherine C. Horton, Shahed Hossain, Irwin Law, Frank van Leth, Guy B. Marks, Hoa Binh Nguyen, Hai Viet Nguyen, Ikushi Onozaki, Maria Imelda D. Quelapio, Alexandra S Richards, Nabila Shaikh, Edine W. Tiemersma, Richard G White, Khalequ Zaman, Frank Cobelens, Rein MGJ Houben

**Author notes:** Corresponding author: Jon C. Emery. **Funding acknowledgements**. JCE, KCH, ASR, NS, and RH have received funding from the European Research Council (ERC) under the Horizon 2020 research and innovation programme (ERC Starting Grant No. 757699). RGW is funded by the Wellcome Trust (218261/Z/19/Z), NIH (1R01AI147321-01), EDTCP (RIA208D-2505B), UK MRC (CCF17-7779 via SET Bloomsbury), ESRC (ES/P008011/1), BMGF (OPP1084276, OPP1135288 & INV-001754), and the WHO (2020/985800-0).

## Abstract

**Background:** Individuals with bacteriologically confirmed pulmonary tuberculosis disease (TB) that do not report symptoms (subclinical TB) represent around half of all prevalent cases of TB, yet their contribution to *Mycobacterium tuberculosis* (*Mtb*) transmission is unknown, especially compared to individuals who report symptoms at time of diagnosis (clinical TB). Relative infectiousness can be approximated by cumulative infections in household contacts, but such data are rare.

**Methods and Findings:** We reviewed the literature to identify studies where surveys of *Mtb* infection were linked to population surveys of TB disease. We collated individual population data for analysis and used literature on the relative durations of subclinical and clinical TB to estimate relative infectiousness through a cumulative hazard model, accounting for sputum-smear status. Relative prevalence of subclinical and clinical disease in high burden settings was used to estimate the contribution of subclinical TB to global *Mtb* transmission.

We collated data on 414 index cases and 789 household contacts from three prevalence surveys (Bangladesh, Philippines, Viet Nam) and one case-finding trial in Viet Nam. The odds ratio of household infection prevalence was 1.2 (0.6-2.3, 95% Confidence Interval). Adjusting for duration of disease, we found a per-unit-time infectiousness of subclinical TB relative to clinical TB of 1.93 (0.62-6.18, 95% Prediction Interval (PrI)). 14 countries across Asia and Africa provided data on relative prevalence of subclinical and clinical TB, suggesting an estimated 68% (27-92%, 95% PrI) of global transmission is from subclinical TB.

**Conclusions:** Our results suggest that subclinical TB contributes substantially to transmission and needs to be diagnosed and treated for effective progress towards TB elimination.

## Introduction

An estimated 1.5-million people died from tuberculosis (TB) disease in 2020, and TB is on course to retake its position as the largest cause of death by a single infectious agent [1]. Fuelled by ongoing transmission through exhaled or expectorated *Mycobacterium tuberculosis* (*Mtb*) bacteria, TB incidence is declining at a rate of 1-2% per annum, which is too slow given the risk and scale of mortality [1,2], lifelong impairment [3,4], poverty [5] and macro-economic consequences [6]. Problematically, most *Mtb* transmission in high incidence settings remains unaccounted for [3], with less than one-in-ten occurrences of TB explained by transmission from a known contact [7].

In recent decades the prevailing paradigm in TB policy held that symptoms and infectiousness commence simultaneously as part of ‘active disease’ [8–10]. As a consequence, a policy of passive case-finding [11], in which individuals are expected to attend a health facility with TB-related symptoms before receiving diagnosis and treatment, was relied upon to prevent deaths from TB, which it has [1,12], and reduce incidence by interrupting transmission, which it has not [1].

Over the last decade, this classic paradigm of TB has been increasingly challenged [13–15]. One important advance has been the finding in population surveys that not all individuals identified with bacteriologically-confirmed TB report having symptoms such as cough at the time of screening for TB [16,17]. As such we can make a distinction between clinical and subclinical TB, where subclinical TB (sometimes referred to as ‘asymptomatic’ [18] or ‘early’ TB [19]) refers to individuals who have detectable *Mtb* bacteria in their sputum but do not experience, or are not aware of symptoms [10]. In contrast, individuals with clinical TB disease report symptoms.

Empirical data have shown that bacteriological state (i.e. whether *Mtb* is detectable in pulmonary secretions) is a strong predictor of the potential for transmission. For example, molecular epidemiological studies show that sputum smear-positive individuals (i.e. *Mtb* detected via microscopy) are 3-6 times more likely to be sources for TB disease in contacts compared to smear-negative individuals [20–22]. Surveys of *Mtb* infection prevalence in household contacts provide similar values [23]. These studies focussed on passively diagnosed individuals with clinical disease. It is however increasingly clear that the presence of respiratory symptoms, such as a persistent cough, is not required for the exhalation of potentially *Mtb*-containing aerosols [24–27]. Indeed, whilst recent empirical studies have suggested that tidal breathing may contribute significantly to *Mtb* transmission [27], a disconnect exists between the exhalation of infectious aerosols and symptoms [28] or cough frequency [29] in TB patients. This supports the hypothesis that subclinical disease can contribute, potentially substantially, to transmission [10,19,30].

A recent review found that about half of prevalent bacteriologically-confirmed pulmonary TB disease is subclinical [18] and it is becoming increasingly apparent that subclinical TB can persist for a long period without progressing to clinical disease [31,32]. As individuals with subclinical TB will not be identified by current passive case-finding strategies, they will continue to contact susceptible individuals and, if infectious, transmit throughout their subclinical phase. It is therefore possible that those with subclinical TB may be a major contributor to ongoing, and unaccounted for, *Mtb* transmission. If this is the case, and if the ambitious goal to end TB as a global health problem by 2035 is to be met [33], TB policy needs to shift away from solely focussing on symptom-dependent case-finding (e.g. patient initiated passive case-finding) toward strategies that are symptom-independent.

To motivate and inform such a shift in research and policy priorities, two key questions that to date remain unanswered must be addressed. Firstly, how infectious are individuals with subclinical TB compared to those with clinical TB per unit time and, secondly, what is their contribution to overall transmission in the current TB epidemic?

In TB, data sources on the transmission potential from sputum smear-negative individuals relative to smear-positive (e.g. molecular epidemiological studies [20–22]) have often been directly interpreted as relative infectiousness, which is incorrect [34]. Instead of representing the metric of interest, which is the potential for transmission per unit time for a particular group relative to a reference group (i.e. relative infectiousness), these data actually provided a relative estimate of cumulative exposure (as acknowledged by these studies’ authors [20–22]). Cumulative exposure is a composite of relative infectiousness per unit time and disease duration (technically duration of infectiousness), which until now have been unavailable and can be hard to disentangle from each other [34].

In this work we look to overcome these challenges by harnessing increased understanding of the natural history and prevalence of subclinical TB and re-analysing data from existing population studies.

## Methods

### Data

To estimate the infectiousness of subclinical TB relative to clinical TB we considered studies in which *Mtb* infection surveys were performed amongst household contacts of culture and/or Nucleic Acid Amplification Test (NAAT) confirmed cases where data on their symptom and sputum smear status at the time of diagnosis was available. We considered only studies in which households with no index case were also surveyed for *Mtb* infection, as a measure of the background rate of infection.

Such studies identified index cases using symptom-independent screening, either via a TB prevalence survey (in which all individuals are screened with a chest X-ray [35]) or community-wide active case-finding amongst a representative sample of a target population. Subclinical and clinical index cases were defined as being culture and/or NAAT positive and responding negatively or positively to an initial symptom screening, respectively. Households with a single subclinical or clinical index case were defined as subclinical and clinical households, respectively. Such households were then stratified by the sputum smear-status of the index case at the time of diagnosis. Background households were defined as having no index case. Finally, *Mtb* infection surveys were performed amongst all households, providing the prevalence of infection amongst each household type.

We reviewed the literature for household contact studies that measure *Mtb* infection via Tuberculin Skin Test (TST) or Interferon-Gamma Release Assay (IGRA) as an outcome and provide sufficient information to stratify households by symptom and sputum smear status, including households with no index case (see **Supplementary Materials** for the detailed search strategy). Individual, patient-level data from each of these studies were analysed to provide the prevalence of infection amongst each household type (see **Supplementary Materials** for detailed data analysis). This data is presented in **Supplementary Table 1**. Odds ratios for infection in members of a household with a sputum smear-positive versus a smear-negative index case (irrespective of symptoms) and in members of a household with a clinical versus subclinical index case (irrespective of sputum smear-status) were calculated.

### Cumulative hazard model

To estimate the infectiousness of subclinical TB per unit time relative to clinical TB, we fitted a cumulative hazard model of infection to the prevalence of infection amongst each household type for each study separately using the data described above.

For each study, household contacts were pooled into five cohorts: background; subclinical and sputum smear-negative; subclinical and sputum smear-positive; clinical and sputum smear-negative; clinical and sputum smear-positive. It was assumed that each cohort is exposed to the same background hazard, reflecting the force of infection from outside the household. It was then assumed that all cohorts except the background were exposed to an additional hazard, reflecting the force of infection from the cohort’s respective index cases.

The final prevalence of infection in each cohort will then depend on the background cumulative hazard *Λ*_*B*_ and an additional cumulative hazard *Λ*_*I*_ specific to each household type *I* (see **Supplementary Materials** for model equations). We use the cumulative hazard from clinical (*C*), smear-positive (+) index cases as a benchmark with which to define the cumulative hazards from the remaining index case types. We assume that being subclinical (*S*) or smear-negative (*-*) have separate, multiplicative effects, such that:

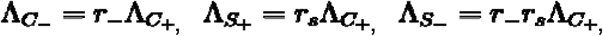

where *r*_*s*_ and r_-_ are the subclinical and sputum smear-negative relative cumulative hazards, respectively.

### Model fitting

The model described above was fitted to the prevalence of infection in each of the five household types for each study separately. Fitting was performed in a Bayesian framework using Markov-Chain Monte-Carlo methods (see **Supplementary Materials** for further details of model fitting). We report median and 95% equal-tailed Posterior Intervals (PoIs).

### Relative infectiousness of subclinical TB

To infer the infectiousness of subclinical TB per unit time relative to clinical TB from our posterior estimate for the subclinical relative cumulative hazard *r*_*s*_, information on the duration of subclinical TB relative to clinical TB was required (see **Supplementary Materials** for details of inferring relative infectiousness from estimated relative cumulative hazard).

As such, we used the results from a recent study that estimated the durations of subclinical and clinical TB using a Bayesian analysis of prevalence and notification data [32]. With the result that the subclinical phase represented between 27% and 63% of the time as a prevalent case, we used a duration of subclinical TB relative to clinical TB of 0.8 (0.4-1.7, 95% PoI). We assumed that there was no difference in duration for sputum smear-negative versus smear-positive TB.

### Subclinical versus clinical TB: prevalence and bacteriological indicators

To estimate the proportion of overall transmission from subclinical TB, we first estimated the proportion of prevalent TB that is subclinical as well as the proportion of prevalent subclinical and clinical TB that is smear-positive.

We began with a recent review of TB prevalence surveys in Asia and Africa [18] (see **Supplementary Materials** for details of the search strategy). Such surveys generally performed an initial screening using both a questionnaire, which includes questions about recent symptoms typical of TB, as well a chest radiograph. Those screening positive from either method were then tested via culture and/or NAAT. A sputum smear-test was often additionally performed.

We reviewed the surveys in [18] and, for each survey where sufficient information was available, extracted the number of culture and/or NAAT confirmed cases of TB, stratified by both symptom status at initial screening and sputum smear status (see **Supplementary Materials** for detailed data analysis). Extracted data can be found in the **Supplementary Table 2**. We defined subclinical and clinical TB as being culture and/or NAAT positive and responding negatively or positively to an initial symptom screen, respectively, consistent with the definitions for subclinical and clinical index cases in the previous section. The most common screening question was a productive cough of greater than 2-weeks duration, although other diagnostic algorithms were included.

For each survey, we calculated the proportion of prevalent TB that is subclinical (*P*^*S*^_*TB*_) as well as the proportion of prevalent subclinical and clinical TB that is smear-positive (*P*^*+*^_*S*_ and *P*^*+*^_*C*_, respectively).

We performed univariate, random-effects meta-analyses on *P*^*S*^_*TB*_, *P*^*+*^_*S*_ and *P*^*+*^_*C*_. We meta-analysed the inverse logit transformed variables, before transforming the results back to proportions and presenting a central estimate and 95% prediction interval for each variable.

### The contribution of subclinical TB to transmission

To estimate the contribution of subclinical TB to transmission, we applied our estimates of relative infectiousness to the prevalence surveys that reported the required data by symptom and smear status (see **Supplementary Materials** for further details).

All analyses were conducted using R version 4.0.3 [36]. Bayesian fitting was performed in Stan version 2.21.0 [37] using RStan [38] as an interface.

### Sensitivity analyses

#### Sensitivity analysis 1

Given the different designs of the Bangladesh (2007) prevalence survey [39] (which provided only sputum smear-positive index cases) and the active case-finding trial in Viet Nam (2017) [40] (which provided index cases via repeated case-finding related screening and prevalence surveys), the above analysis was repeated omitting these studies.

#### Sensitivity analysis 2

As a sensitivity the above analysis was repeated with an alternative estimate for the relative duration of subclinical TB versus clinical TB, using instead data from a recent systematic review and data synthesis study [31] and a simple competing risk model (see **Supplementary Materials** for further details).

## Results

### Estimating the relative infectiousness of subclinical TB

Four studies were included for analysis: three prevalence surveys of TB disease with associated *Mtb* infection surveys in Viet Nam (2007) [41], Bangladesh (2007) [39] and the Philippines (1997) [42] and a community-wide active case-finding trial in Viet Nam (2017) [40].

Odds ratios (ORs) for infection in members of a household with a sputum smear-positive versus a smear-negative index case (irrespective of symptoms) are shown in **Figure 1A**, where the Bangladesh prevalence survey is omitted as this study only included smear-positive individuals. A mixed-effects meta-analysis across studies provides OR = 2.3 (1.3-3.9, 95% confidence interval (CI)), which is in line with previous estimates [23]. **Figure 1B** shows the odds ratio for infection in members of a household with a clinical versus subclinical index case (irrespective of sputum smear-status), based on the result of their symptom screen at the time of diagnosis. In contrast to the analysis by smear status, no evidence for a difference in cumulative infection was found by symptom status, with OR = 1.2 (0.6-2.3, 95% CI).

**Figure 1:**
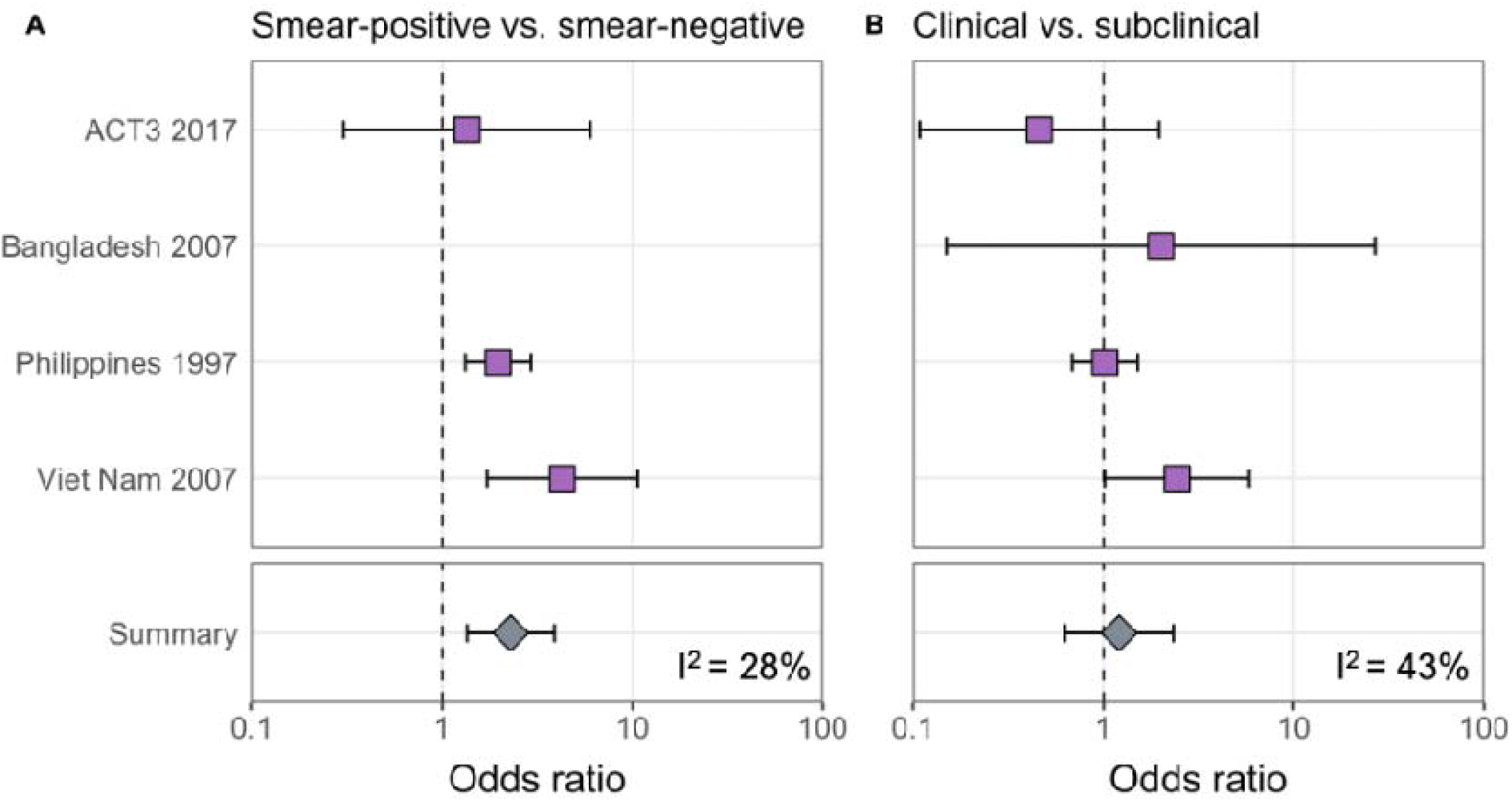
Odds ratios for infection in members of a household with a sputum smear-positive versus a smear-negative index case (irrespective of symptoms) (**A**) and in members of a household with a clinical versus a subclinical index case (irrespective of sputum smear-status) (**B**). Illustrated are central estimates and 95% confidence intervals for each study separately and the results of a mixed-effects meta-analysis. Results for sputum smear status are omitted for Bangladesh as the survey considered only sputum smear-positive individuals

The estimated infectiousness of subclinical TB per unit time relative to clinical TB is shown in **Figure 2A**, both for each study separately as well as the mixed-effects meta-analysed result across studies of 1.93 (0.62-6.18, 95% Prediction Interval (PrI)). **Figure 2B** shows the analogous results for the infectiousness per unit time of sputum smear-negative versus smear-positive TB, with a summary value of 0.26 (0.03-2.47, 95% PrI). Detailed model results are shown in **Supplementary Table 4** and **Supplementary Figures 2-5**.

**Figure 2:**
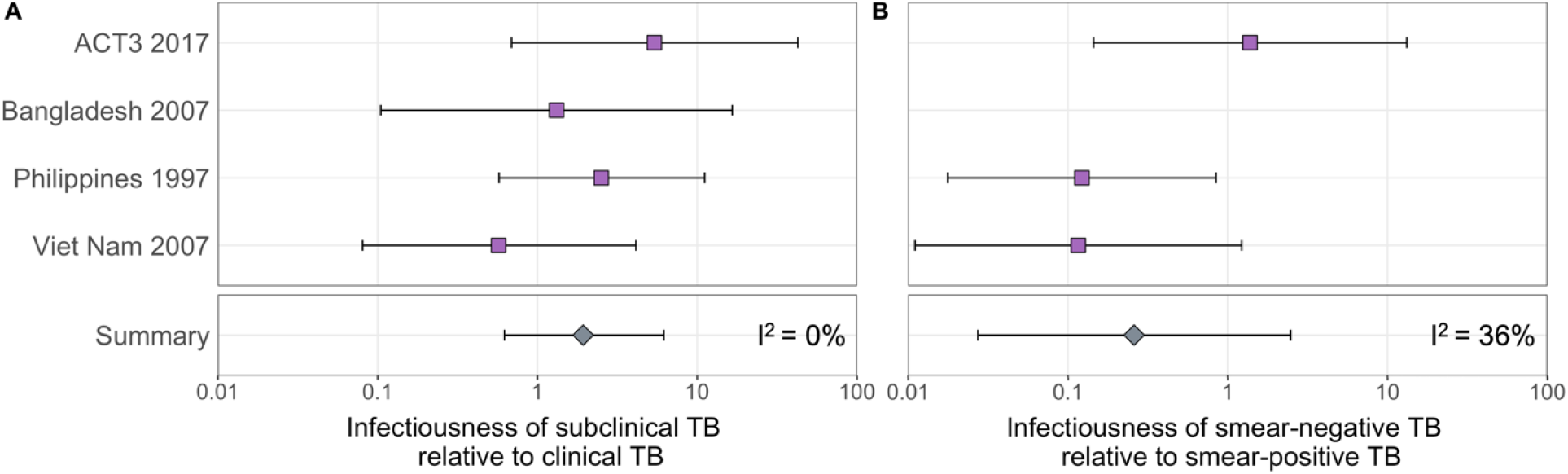
The estimated infectiousness of subclinical TB per unit time relative to clinical TB (**A**) and sputum smear-negative TB relative to smear-positive TB (**B**). Illustrated are the median and 95% confidence intervals for each study separately and the median and 95% prediction interval results from mixed-effects meta-analyses across studies with an associated measure of heterogeneity (I^2^).

### Prevalence and bacteriological indicators for subclinical and clinical TB

Data from 15 prevalence surveys where the proportion of subclinical and clinical TB was reported by sputum smear-status were included, detailed in **Supplementary Table 2**. These represented a range of high TB burden countries in Africa (n=5) and Asia (n=9, with two surveys in Viet Nam). In this subset, the overall proportion of prevalent TB that is subclinical was 58% (29-82%, 95% PrI), whilst the proportion smear-positive was 33% (18-52%, 95% PrI) for subclinical TB and 53% (25-80%, 95% PrI) for clinical TB. Detailed results for each variable are shown in **Figure 3A-C**.

**Figure 3:**
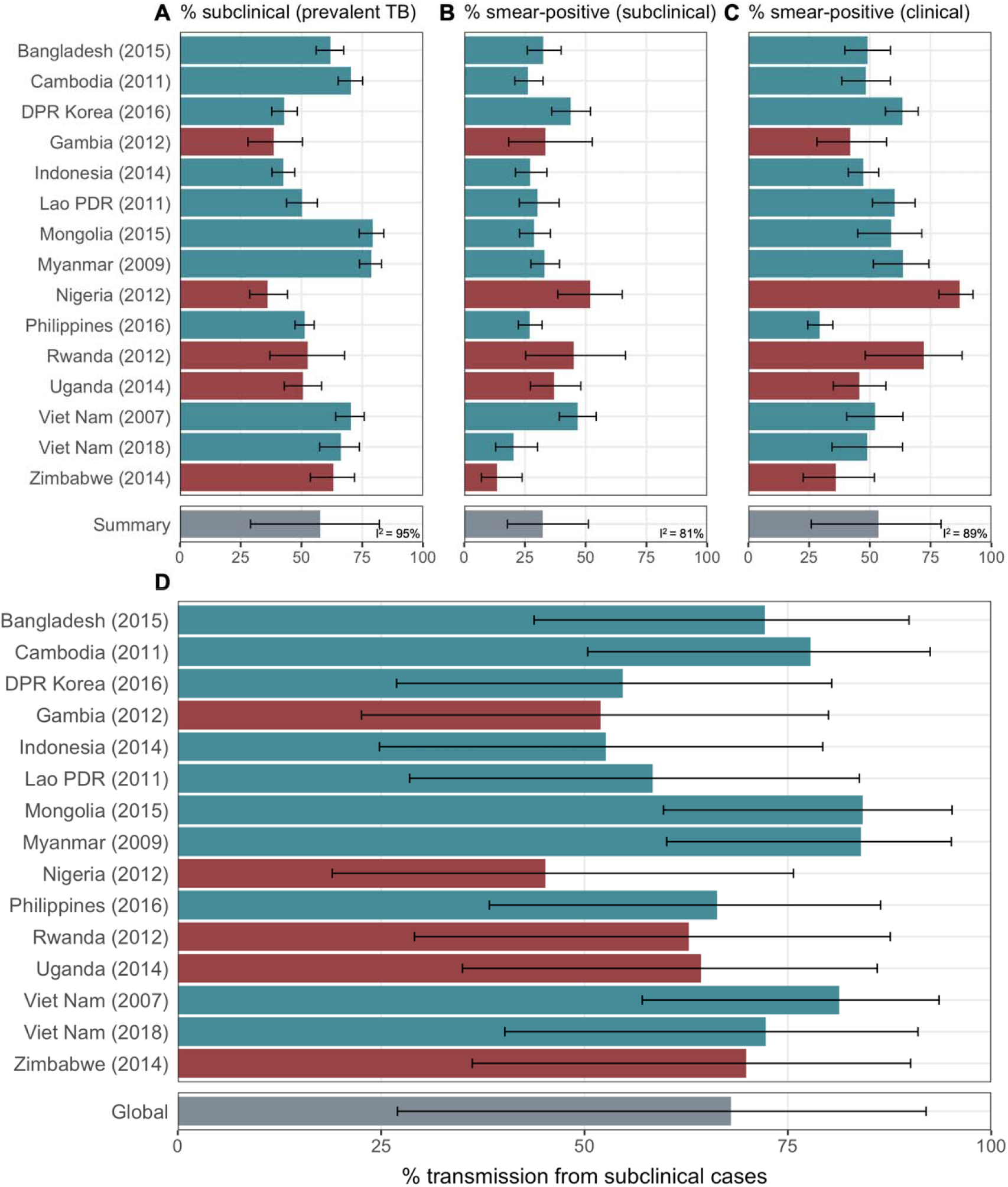
The proportion of prevalent TB that is subclinical (**A**), the proportion of subclinical TB that is smear-positive (**B**) and the proportion of clinical TB that is smear-positive (**C**) using data from prevalence surveys in Africa (red) and Asia (teal). Illustrated are median and 95% confidence intervals for each study separately and the median and 95% prediction intervals from mixed-effects meta-analyses across studies with an associated measure of heterogeneity (I^2^). Also shown is the estimated proportion of transmission from subclinical TB at the time of and in the location of each of the prevalence surveys in Africa and Asia (**D**). Illustrated is the median and 95% prediction intervals for each study separately as well as the global value. DPR = Democratic People’s Republic; PDR = People’s Democratic Republic.

### The contribution of subclinical TB to transmission - global and country levels

We quantified the contribution of subclinical TB to ongoing *Mtb* transmission by combining the estimates for the infectiousness of subclinical TB per unit time relative to clinical TB (**Figure 2A**), the infectiousness of sputum smear-negative TB relative to smear-positive TB (**Figure 2B**), the proportion of prevalent TB that is subclinical and the proportion of subclinical and clinical TB that is sputum smear-positive (**Figure 3A-C**). The 14 included countries are a reasonable reflection of the geography and epidemiological characteristics of high TB burden countries in the WHO African, South-East Asia and Western Pacific regions, which together represent around 85% of current global TB incidence [1]. As such we used a summary value for the included surveys as a global estimate.

**Figure 3D** shows the results by country and globally, where 68% (27-92%, 95% PrI) of global *Mtb* transmission is estimated to come from prevalent subclinical TB, ranging from 45% (19%-76%, 95% PrI) in Nigeria to 84% (60%-95%, 95% PrI) in Mongolia.

### Sensitivity analyses

#### Sensitivity analysis 1

The above analysis was repeated excluding two studies with methodologies that differed from the remaining two: the Bangladesh (2007) prevalence survey [39] (which provided sputum smear-positive index cases only); and the active case-finding trial in Viet Nam (ACT3 (2017)) [40] (which provided index cases via repeated screening related to case-finding as well as prevalence surveys). Affected results are shown in **Supplementary Figure 6**. The infectiousness of subclinical TB per unit time relative to clinical TB decreased to 1.39 (0.17-11.2, 95% PrI) and the infectiousness of sputum smear-negative TB relative to smear-positive TB decreased to 0.12 (0.03-0.53, 95% PrI), with corresponding values of 57% (10-94%, 95% PrI) of global transmission from subclinical TB, ranging from 34% (6%-81%, 95% PrI) in Nigeria to 76% (28%-97%, 95% PrI) in Mongolia.

#### Sensitivity analysis 2

As a sensitivity the above analysis was repeated using an alternative estimate for the relative duration of subclinical TB versus clinical TB of 0.72 (0.60-0.89, 95% PoI), from [32]. Affected results are shown in **Supplementary Figure 7**. The infectiousness of subclinical TB per unit time relative to clinical TB increased to 2.19 (0.91-5.26, 95% PrI), with corresponding values of 71% (32-92%, 95% PrI) of global transmission from subclinical TB, ranging from 48% (25-74%, 95% PrI) in Nigeria to 86% (68-95%, 95% PrI) in Mongolia.

## Discussion

By fitting a cumulative hazard model of infection to prevalence data amongst household contacts of subclinical and clinical index cases, we were able to provide quantitative estimates for the relative infectiousness per unit time of subclinical TB and its contribution to ongoing *Mtb* transmission. Despite wide uncertainty intervals, the raw data, as well as the results of our analysis, do not suggest subclinical TB is substantially less infectious than clinical TB. Given the high prevalence of subclinical TB found in surveys [18], it is therefore likely that subclinical TB contributes substantially to ongoing *Mtb* transmission in high burden settings.

There are no other estimates for the infectiousness of subclinical TB relative to clinical TB in the literature with which to compare our results, although a recent small study from Uganda found no evidence of a difference in cumulative infection rates in household contacts of patients who did or did not report symptoms [43]. Previous hypothetical modelling of subclinical TB used assumed values for relative infectiousness that are in keeping with our estimated range [30,44]. Recent work on SARS-CoV-2 and malaria have similarly shown how ‘asymptomatic’ or ‘subpatent’ infections can be important drivers of transmission [45–47].

Whilst we have presented a novel approach to investigating transmission from individuals with subclinical TB using pre-existing data, limitations in our methodology remain. Identifying relative infectiousness is challenging. Our estimates rely on studies which screened a minimum of 252,000 individuals for bacteriologically confirmed TB disease, and 63,000 individuals for *Mtb* infection. Even at this scale, the small number of diagnosed cases of TB still leads to substantial uncertainty, highlighting the challenge faced by single studies to estimate such values [40,43].

Although infection studies in household contacts have provided a novel window into transmission from subclinical individuals, it is not possible to establish a transmission link between presumed index cases and infections amongst household contacts using molecular methods [34]. Whilst our model does use a background rate of infection as a baseline from which to estimate the additional force of infection from presumed index cases within the household, there remains the residual risk that certain household types may systematically contain more or less infections from transmission outside the household than on average.

Our cumulative hazard model assumed that index cases only ever had the disease type they were diagnosed with during screening. Instead, it is more likely that individuals will fluctuate between being subclinical and clinical [31]. The impact such additional dynamics would have on our results remains uncertain and would depend on the detailed model of tuberculosis natural history assumed.

We estimated the contribution of subclinical TB to transmission at the population level, including transmission outside the household, using information on relative infectiousness inferred from household contact studies. A more refined estimate may need to take additional factors into account. For example it is likely that, whilst inside the household the contact rates for subclinical and clinical individuals are likely to be similar, contact with individuals outside the household may differ [48].

Meta-analyses were used to provide ranges for several quantities of interest. Whilst the heterogeneity for the relative infectiousness of subclinical and smear-negative TB were low (*I*^*2*^ = 0% and *I*^*2*^ = 36%, respectively), the heterogeneity for the proportion of prevalent TB that is subclinical and the proportions of subclinical and clinical TB that are smear-positive were high (*I*^*2*^ = 95%, *I*^*2*^ = 81% and *I*^*2*^ = 89%, respectively). As such, we have used the more conservative prediction interval (as opposed to credible interval) to reflect this heterogeneity in the final results [49,50].

Finally, our data are from populations with a low prevalence of HIV co-infection and the HIV status of individuals with TB was mostly unavailable, making a sub-analysis by HIV and antiretroviral (ART) status impossible. Whilst the subclinical TB presentation is likely affected by HIV in terms of duration and prevalence, it is unknown whether or by how much the relative differences in duration and prevalence between subclinical and clinical TB also change [18,51]. If they exist, any such differences by HIV-coinfection status are likely to be reduced by effective viral suppression, which an estimated two-thirds of people living with HIV have achieved [52]. So while it remains highly valuable to accumulate additional relevant data [43], we feel our main findings are broadly robust to this limitation.

Our observation that reported symptoms are a poor proxy for infectiousness fits with historical and contemporary observations that symptom-independent TB screening and treatment policies can reduce TB burden at higher rates than usually seen under DOTS [40,53]. This is in keeping with increasing data showing that symptoms, in particular the classic TB symptom of cough, are not closely correlated to the amount of *Mtb* exhaled [28,29] and observations of other pathogens, including SARS-CoV-2 infection [46,47,54].

Whilst earlier diagnosis (i.e. before symptom onset) will likely bring individual-level benefits in terms of mortality and extent of post-TB sequelae [55], the question of potential population benefits has hampered decisions from policy makers and funders on whether to invest resources in technologies and strategies that can identify subclinical TB. Our results suggest that a non-trivial proportion of all transmission would likely be unaffected by strategies that are insensitive to subclinical TB.

Natural next steps could include adding subclinical TB to the planned update of WHO case definitions and to develop Target Product Profiles for tools that can detect all infectious TB, regardless of whether individuals are experiencing or aware of symptoms, and to critically evaluate such tools in interventions for their impact on *Mtb* transmission and cost-effectiveness. While symptom-independent tools exist for screening [29,40,56–59], specificity, costs and logistics remain an obstacle. In addition, individuals are usually required to produce sputum as part of a confirmatory test, which around half of eligible adults in the general population are unable to do [40]. Screening or diagnostic technologies that are symptom-as well as sputum-independent, while remaining low-tech and low-cost, remain the goal. Indeed, the advent of bio-aerosol measurements in TB may uncover additional infectious individuals whose sputum-based bacteriological test is negative, although these tools require validation in larger populations [29,59,60]. Any bio-aerosol positive, sputum-negative individuals are more likely to be subclinical and as such would mean we underestimated the contribution of subclinical TB to global *Mtb* transmission, even if their relative infectiousness may be lower than sputum-positive TB. As new diagnostic approaches are developed to capture the spectrum of TB disease, policy makers will need to decide on how to treat subclinical TB. In treatment, as in diagnosis, it is key that more tailored approaches are developed and tested, so as to prevent over- or undertreatment of individuals with subclinical TB [61].

## Conclusion

Subclinical TB likely contributes substantially to transmission in high burden settings. If we are to meet EndTB targets for TB elimination, the TB community needs to develop technologies and strategies beyond passive case finding to address subclinical TB.

## Data Availability

Analysis scripts and replication data are available online at https://github.com/epidemery/subclinical_transmission.

## Data availability

Replication data are available at https://github.com/epidemery/subclinical_transmission.

## Code availability

Analysis scripts are available at https://github.com/epidemery/subclinical_transmission.

## Acknowledgements

The authors would like to acknowledge the work of Dr Thelma Tupasi who led the Philippines (2007) prevalence survey and founded the Tropical Disease Foundation (Philippines), who sadly passed away in 2019.

## Author contributions

RH conceived the study; JE, RH, PD and FC designed the study; JE, RH, BF, FG and HVN performed data processing; JE led the analysis with support from PD and RH; JE and NS produced the visualisations; JE wrote the first draft; All contributed to writing and reviewed the final draft.

