## Supplementary materials for "Estimating the contribution of subclinical tuberculosis disease to transmission – an individual patient data analysis from prevalence surveys"

### Supplementary methods

#### *Data*

*Search strategy*

We sought studies in which *Mtb* infection surveys were performed amongst household contacts of bacteriologically-confirmed index cases, where data on their symptom and sputum smear-status at the time of diagnosis was available.

We began with a recent systematic review of population-based TB prevalence surveys completed since 1990, with reports or articles publicly available through August 2019 [1]. Surveys were included if both a symptom screening interview and X-ray were performed on all eligible participants and if surveys reported the proportion of bacteriologically-confirmed cases by screening modality as well as the proportion of bacteriologically confirmed cases that were negative on symptom screening (see [1] for full details of the review process). We then reviewed the reports of the 28 national and subnational TB prevalence surveys included for quantitative analysis in [1] and identified 3 such studies that were conducted alongside *Mtb* infection surveys amongst household contacts: Viet Nam (2007) [2], Bangladesh (2007) [3] and the Philippines (1997) [4]. Authors of these studies and affiliated institutions were then invited to collaborate using original, individual-level data and all accepted.

In addition to prevalence surveys we also considered active case-finding studies with associated household infection surveys with which to measure any resultant impact on transmission. A non-systematic review of the literature identified one such study in Viet Nam: ACT3 (2017) [5]. Again the authors of this study and affiliated institutions were invited to collaborate with original, individual-level data and accepted.

*Data analysis*

Index cases were identified in the three prevalence surveys (Viet Nam (2007) [2], Bangladesh (2007) [3] and the Philippines (1997) [4]) via culture and/or NAAT and defined as subclinical or clinical depending on whether they responded negatively or positively to an initial symptom screening, respectively. Index cases were further stratified by their sputum smear-status at the time of diagnosis.

Linked records were then used to stratify participants of the associated *Mtb* infection survey into different household types depending on the status of the index case: background (no index case); subclinical and sputum smear-negative; subclinical and sputum smear-positive; clinical and sputum smear-negative; clinical and sputum smear-positive. For each household type the total number of contacts and number of TST or IGRA-positive contacts was extracted, shown in **Supplementary Table 1**.

Index cases were identified in ACT3 [5] either through routine passive case-finding, in any of the TB screening rounds as part of active case-finding or in the TB prevalence surveys used to measure the impact of such screening. Those identified through passive case-finding were designated clinical, whilst those identified either through screening or the prevalence surveys were stratified as subclinical or clinical depending on whether they responded negatively or positively to an initial symptom screening, respectively. Index cases were further stratified by their sputum smear-status at the time of diagnosis.

The same approach to that described above was then used to find the total number of household contacts and the number of TST or IGRA-positive contacts for each household type, also shown in **Supplementary Table 1**.

| Study | Background | | Subclinical | | | | Clinical | | | |
|---|---|---|---|---|---|---|---|---|---|---|
| | | | Smear-negative | | Smear-positive | | Smear-negative | | Smear-positive | |
| | Infected | Contacts | Infected | Contacts | Infected | Contacts | Infected | Contacts | Infected | Contacts |
| ACT3 2017 [5] | 128 | 2893 | 2 | 8 | 2 | 10 | 1 | 16 | 4 | 27 |
| Bangladesh 2007 [3] | 702 | 17566 | *NA* | *NA* | 1 | 5 | *NA* | *NA* | 3 | 9 |
| Philippines 1997 [4] | 3823 | 20259 | 48 | 227 | 32 | 82 | 23 | 108 | 34 | 109 |
| Vietnam 2007 [6] | 1556 | 21298 | 3 | 59 | 5 | 28 | 4 | 42 | 16 | 59 |

**Supplementary Table 1:** Summary of the relevant data from studies in which *Mtb* infection surveys were performed amongst household contacts of culture and/or NAAT confirmed cases where information on their symptom and sputum smear-status at the time of diagnosis was available. *Infected = Number of TST or IGRA-positive household contacts; Contacts = Number of household contacts with a TST or IGRA result. NA = Not applicable*

***Cumulative hazard model***

*Model equations*

The prevalence of infection in background households (i.e. with no index case) is given by:

$$P_B = 1 - e^{-\Lambda_B}$$

where $\Lambda_B$ is the cumulative hazard from the background, representing transmission outside the household. The prevalence of infection in households with an index case is given by:

$$P_I = 1 - e^{-\Lambda_B} e^{-\Lambda_I}$$

where $\Lambda_I$ is the cumulative hazard from index case type *I = subclinical and smear-negative (S-); subclinical and smear-positive (S+); clinical and smear-negative (C-); clinical and smear-positive (C+)*

The cumulative hazard from clinical and smear-positive index cases is used as a benchmark to define the cumulative hazards from the remaining index case types. We assume that being subclinical or smear-negative have separate, multiplicative effects, such that:

$$\Lambda_{C_-} = r_-\Lambda_{C_+}, \quad \Lambda_{S_+} = r_s\Lambda_{C_+}, \quad \Lambda_{S_-} = r_-r_s\Lambda_{C_+},$$

where $r_s$ and $r_-$ are the subclinical and sputum smear-negative relative cumulative hazards, respectively.

#### *Model fitting*

The model was fitted to the prevalence of infection in each of the five household types for each study separately. Fitting was performed in a Bayesian framework using Markov-Chain Monte-Carlo methods. We use binomial distributions for the prevalence in the likelihood and estimate the following parameters: the background cumulative hazard ($\Lambda_B$); the cumulative hazard from clinical, smear-positive index cases ($\Lambda_{C+}$), the subclinical relative cumulative hazard ($r_s$) and the sputum smear-negative relative cumulative hazard ($r_-$). We use truncated gamma and normal distributions as weak priors:

$\Lambda_B$ ~ gamma(alpha = 2, beta = 20),
$\Lambda_{C+}$ ~ normal(mu = 0.5, sigma = 20),
$r_s$ ~ normal(mu = 1, sigma = 20),
$r_-$ ~ normal(mu = 0.2, sigma = 20).

A total of 50,000 iterations were performed for each study, the first 25,000 of which were discarded as burn-in. Model fit, trace, correlation and auto-correlation plots were used to ensure model suitability and convergence. We report median and 95% equal-tailed posterior intervals (PoIs).

***Relative infectiousness of subclinical TB***

*Inferring relative infectiousness per unit time from estimated relative cumulative hazard*

Assuming constant hazards, the relative cumulative hazards from index cases will depend on the product of the relative per unit time infectiousness and relative durations of infectiousness. We assume that:

1. Per unit time infectiousness depends on symptom status and sputum smear-status
2. Durations of infectiousness only depend on symptom status

It follows then that:

$$r_s = \alpha_s \gamma_s,$$
$$r_- = \alpha_-,$$

where $\alpha_s$ and $\alpha_-$ are the per unit time infectiousness of subclinical relative to clinical index cases and sputum smear-negative relative to smear positive index cases, respectively, and $\gamma_s$ is the duration of infectiousness for subclinical relative to clinical index cases.

We sampled from the posterior estimate for the subclinical relative cumulative hazard and assumed duration of disease for subclinical index cases relative to clinical index cases, providing a median and 95% equal tailed posterior estimate for the relative infectiousness of subclinical index cases relative to clinical index cases for each study separately. Finally, we provide a summary estimate by mixed-effects meta-analysing the individual estimates across the separate studies.

Since we assumed that there is no difference in duration for sputum smear-negative versus smear-positive TB, our estimate for the smear-negative relative cumulative hazard is also an estimate for the relative infectiousness per unit time of sputum smear-negative TB relative to smear-positive TB. We provide analogous results to those described above for the relative infectiousness of subclinical TB.

***Subclinical versus clinical TB: prevalence and bacteriological indicators***

*Search strategy*

We sought TB prevalence surveys where data on the symptom and sputum smear-status at the time of diagnosis was available for those identified in the survey. We began again with the recent systematic review of population-based TB prevalence surveys [1], all of which included information on the symptom status at the time of diagnosis of those identified in the survey. We again reviewed the reports of the 28 national and subnational TB prevalence surveys included for quantitative analysis in [1] and identified 14 such studies that also included information on the sputum smear-status at the time of diagnosis of those identified in the survey. Data from the second TB prevalence survey in Viet Nam in 2018 [7], which was not included in [1], were additionally included.

*Data analysis*

From the respective survey reports we extracted the symptom threshold used for initial symptom screening, the total number of individuals screened and the number of identified cases that were: subclinical and sputum smear-negative; subclinical and sputum smear-positive; clinical and sputum smear-negative; clinical and sputum smear-positive. Results of the data extraction are shown in **Supplementary Table 2**.

| Survey setting [Ref] | Year | Subclinical Smear neg. | Subclinical Smear pos. | Clinical Smear neg. | Clinical Smear pos. | Number screened | Symptom threshold |
|---|---|---|---|---|---|---|---|
| Viet Nam [7] | 2018 | 67 | 17 | 22 | 21 | 61763 | Cough > 2 weeks |
| Viet Nam [8] | 2007 | 87 | 76 | 33 | 36 | 94179 | Productive cough > 2 weeks |
| Myanmar [9] | 2009 | 164 | 81 | 24 | 42 | 51367 | Any symptom |
| Lao PDR [10] | 2011 | 83 | 36 | 47 | 71 | 39212 | Cough > 2 weeks and/or other |
| Cambodia [11] | 2011 | 163 | 58 | 48 | 45 | 37417 | Cough > 2 weeks and/or other |
| Gambia [12] | 2012 | 18 | 9 | 25 | 18 | 43100 | Cough > 2 weeks and/or other |
| Rwanda [13] | 2012 | 11 | 9 | 5 | 13 | 43128 | Any symptom |
| Nigeria [14] | 2012 | 25 | 27 | 12 | 80 | 44186 | Cough > 2 weeks |
| Indonesia [15] | 2014 | 132 | 49 | 129 | 116 | 67944 | Cough > 2 weeks and/or other |
| Uganda [16] | 2014 | 51 | 30 | 43 | 36 | 41154 | Cough > 2 weeks |
| Zimbabwe [17] | 2014 | 58 | 9 | 25 | 14 | 33736 | Any symptom |
| Bangladesh [18] | 2015 | 116 | 56 | 54 | 52 | 98710 | Cough > 2 weeks and/or other |
| Mongolia [19] | 2015 | 139 | 56 | 21 | 30 | 50309 | Cough > 2 weeks |
| DPR Korea [20] | 2016 | 82 | 64 | 71 | 123 | 60683 | Cough > 2 weeks and/or other |
| Philippines [21] | 2016 | 231 | 85 | 212 | 88 | 46689 | Cough > 2 weeks and/or other |

**Supplementary Table 2:** Data extracted from 15 prevalence where sufficient information on sputum smear-status at the time of diagnosis was available. The 'symptom threshold' used for initial symptom screening is the metric used here to define subclinical (negative) and clinical (positive). *Neg = Negative, Pos = Positive*.

#### *The contribution of subclinical TB to transmission*

We combined our estimates for the relative infectiousness of subclinical TB per unit time relative to clinical TB ($\alpha_S$), the relative infectiousness of sputum smear-negative TB relative to smear-positive TB ($\alpha_-$), the meta-analysed proportion of prevalent TB that is subclinical ($P^S_{TB}$), and the proportion of prevalent subclinical and clinical TB that is smear-positive ($P^+_S$ and $P^+_C$, respectively) to estimate the per unit time contribution of subclinical TB to overall transmission ($P^S_{Tx}$):

$$P^S_{Tx} = \frac{(P^+_S\alpha_s + (1 - P^+_S)\alpha_s\alpha_-)P^S_{TB}}{(P^+_S\alpha_s + (1 - P^+_S)\alpha_s\alpha_-)P^S_{TB} + (P^+_C + (1 - P^+_C)\alpha_-)(1 - P^S_{TB})}.$$

To this end, we used the posterior distributions for $\alpha_S$ and $\alpha_-$ from the earlier model fitting. We also modelled $P^+_S$, $P^+_C$ and $P^S_{TB}$ as normal distributions with means and variances taken from the univariate meta-analysis described above. The expression for the contribution of subclinical TB to overall transmission was then evaluated using $10^7$ samples where we report the median and equal-tailed 95% prediction intervals.

The above was then re-performed on a survey-by-survey basis. Here $P^+_S$, $P^+_C$ and $P^S_{TB}$ were modelled separately for each survey and assumed to be distributed binomially. The distributions used for $\alpha_S$ and $\alpha_-$ remained unchanged.

#### *Sensitivity analyses*

*Sensitivity analysis 2:* We calculate the duration of infectiousness for subclinical relative to clinical index cases using the model and transition values from [22]. The model is shown in **Supplementary Figure 1A** with associated transition values, which are also detailed in **Supplementary Table 3**. Disease durations are given by the inverse sum of all transitions out of subclinical (regression or progression) or clinical disease (regression, diagnosis and treatment or death). We find durations of 5.4 months (4.6-6.7 months, 95% PoI) and 7.5 months (7.0-8.2 months, 95% PoI) for subclinical and clinical TB, respectively (**Supplementary Figure 1B**), giving a relative duration of subclinical versus clinical TB of 0.72 (0.60-0.89, 95% PoI)).

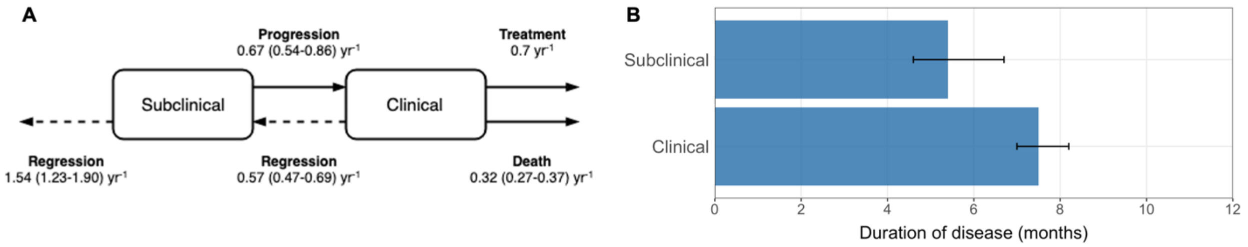


***Supplementary Figure 1:*** *Competing risk model (**A**) with transition rates from* [22] *used to estimate the durations of subclinical and clinical TB (**B**).*

| **Parameter** | **Value (95% posterior interval)** | **Units** |
|---|---|---|
| Regression from subclinical | 1.54 (1.23-1.90) | Per year |
| Progression from subclinical | 0.67 (0.54-0.86) | Per year |
| Regression from clinical | 0.57 (0.47-0.69) | Per year |
| Treatment from clinical | 0.70 | Per year |
| Death from clinical | 0.32 (0.27-0.37) | Per year |

**Supplementary Table 3:** Progression and regression parameter values taken from [22] used to estimate the durations of subclinical and clinical TB using the competing risk method detailed in the main text. See [22] for data sources and methods for estimating the above parameters.

### Supplementary results

#### *Estimating the relative infectiousness of subclinical TB*

*Detailed model results*

Viet Nam

| | n_eff | Rhat | mean | mcse | sd | 2.5% | 50% | 97.5% |
|---|---|---|---|---|---|---|---|---|
| lambda_B | 16,683 | 1 | 0.076 | 0 | 0.002 | 0.072 | 0.076 | 0.08 |
| lambda_Cp | 14,000 | 1 | 0.222 | 0.001 | 0.078 | 0.089 | 0.216 | 0.393 |
| r_s | 11,465 | 1 | 0.653 | 0.006 | 0.61 | 0.05 | 0.524 | 1.988 |
| r_n | 15,677 | 1 | 0.195 | 0.002 | 0.197 | 0.006 | 0.14 | 0.682 |

Philippines

| | n_eff | Rhat | mean | mcse | sd | 2.5% | 50% | 97.5% |
|---|---|---|---|---|---|---|---|---|
| lambda_B | 12,836 | 1 | 0.209 | 0 | 0.003 | 0.203 | 0.209 | 0.216 |
| lambda_Cp | 8,107 | 1 | 0.145 | 0.001 | 0.065 | 0.028 | 0.142 | 0.281 |
| r_s | 4,532 | 1 | 2.644 | 0.044 | 2.94 | 0.683 | 1.91 | 9.7 |
| r_n | 15,022 | 1 | 0.172 | 0.001 | 0.131 | 0.01 | 0.145 | 0.484 |

ACT3

| | n_eff | Rhat | mean | mcse | sd | 2.5% | 50% | 97.5% |
|---|---|---|---|---|---|---|---|---|
| lambda_B | 13,924 | 1 | 0.046 | 0 | 0.004 | 0.038 | 0.046 | 0.054 |
| lambda_Cp | 10,476 | 1 | 0.058 | 0.001 | 0.055 | 0.002 | 0.042 | 0.203 |
| r_s | 9,133 | 1 | 6.843 | 0.074 | 7.052 | 0.612 | 4.406 | 27.192 |
| r_n | 7,628 | 1 | 2.696 | 0.049 | 4.273 | 0.143 | 1.337 | 15.314 |

Bangladesh

| | n_eff | Rhat | mean | mcse | sd | 2.5% | 50% | 97.5% |
|---|---|---|---|---|---|---|---|---|
| lambda_B | 10,804 | 1 | 0.041 | 0 | 0.002 | 0.038 | 0.041 | 0.044 |
| lambda_Cp | 11,415 | 1 | 0.349 | 0.002 | 0.24 | 0.037 | 0.297 | 0.944 |
| r_s | 7,640 | 1 | 2.101 | 0.036 | 3.174 | 0.076 | 1.113 | 10.723 |

**Supplementary Table 4:** Posterior summary statistics for each model. Shown are: the effective sample size (n_eff); the 'R hat' statistic (Rhat); sample mean (mean); Monte Carlo Standard Error (mcse); sample standard deviation (sd); and sample quantiles (2.5%, 50%, 97.5%).

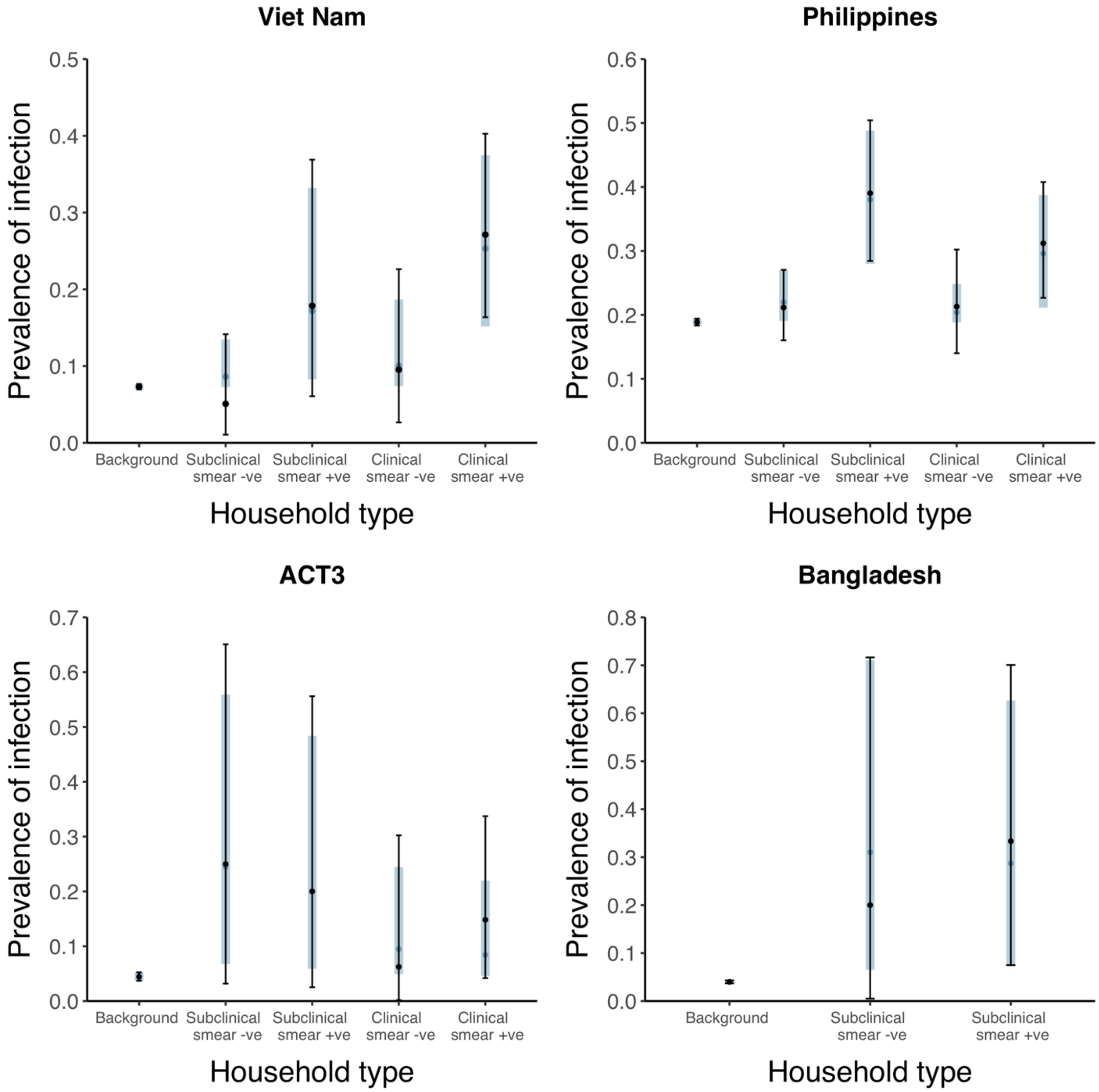


**Supplementary Figure 2:** Model fits for each model. Shown are prevalence of infection in members of households with different index case types (background, subclinical and smear-negative, subclinical and smear-positive, clinical and smear-negative, clinical and smear-positive). Error bars show median and 95% credible intervals. Shaded regions show posterior median and 95% posterior intervals. +*ve = positive, -ve = negative*.

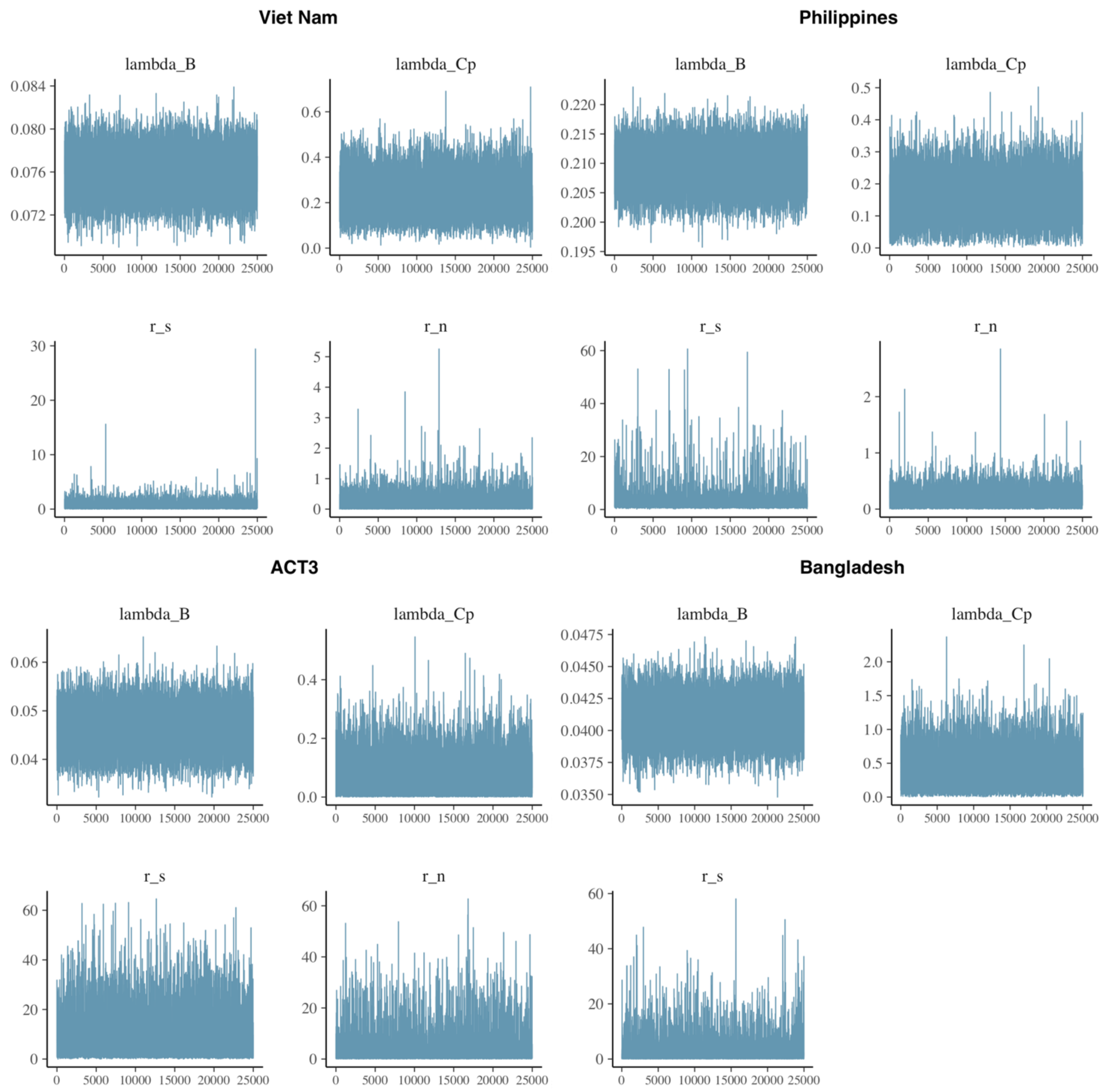


**Supplementary Figure 3:** Trace plots for each model.

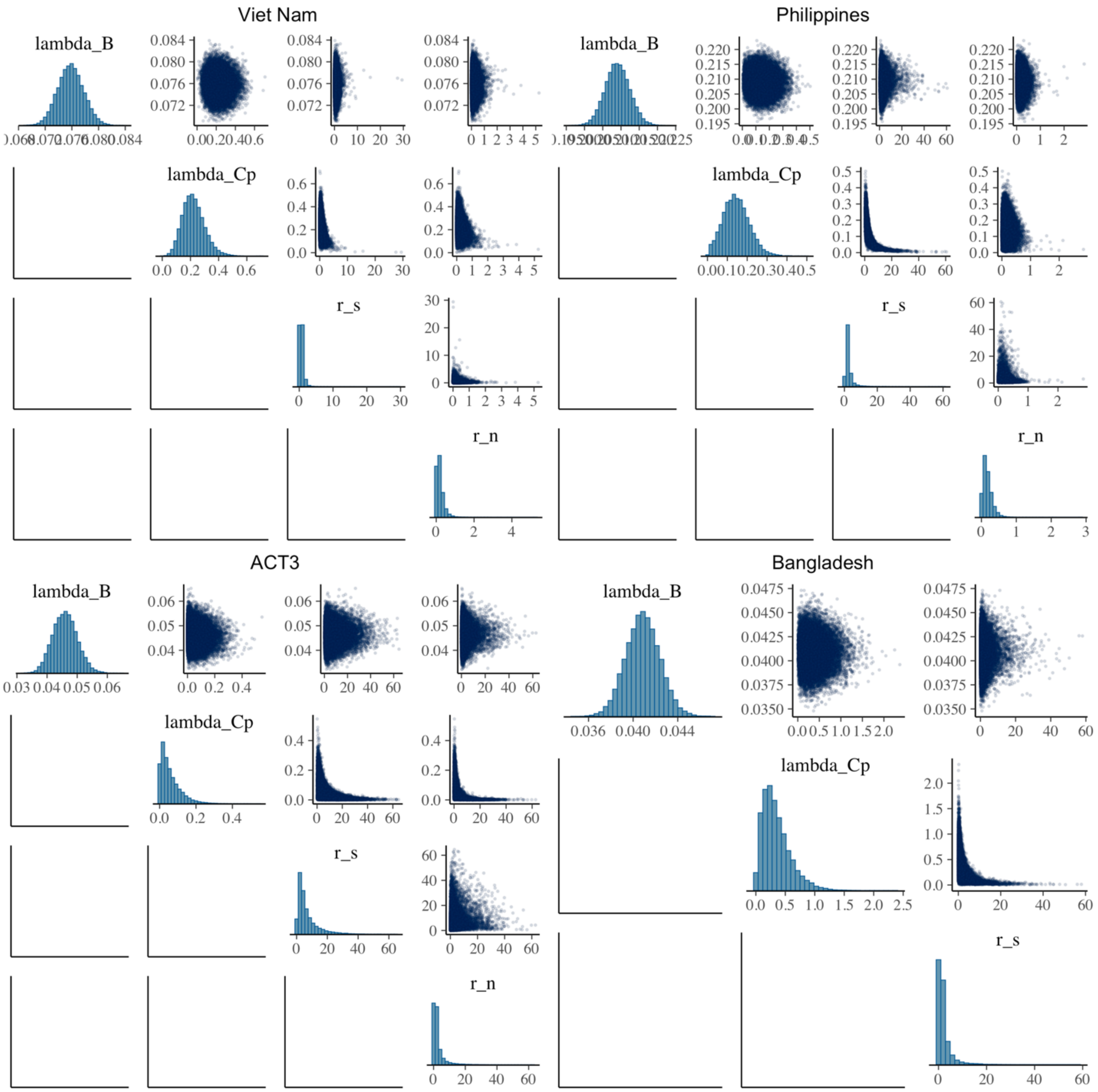


**Supplementary Figure 4:** Correlation plots for each model.

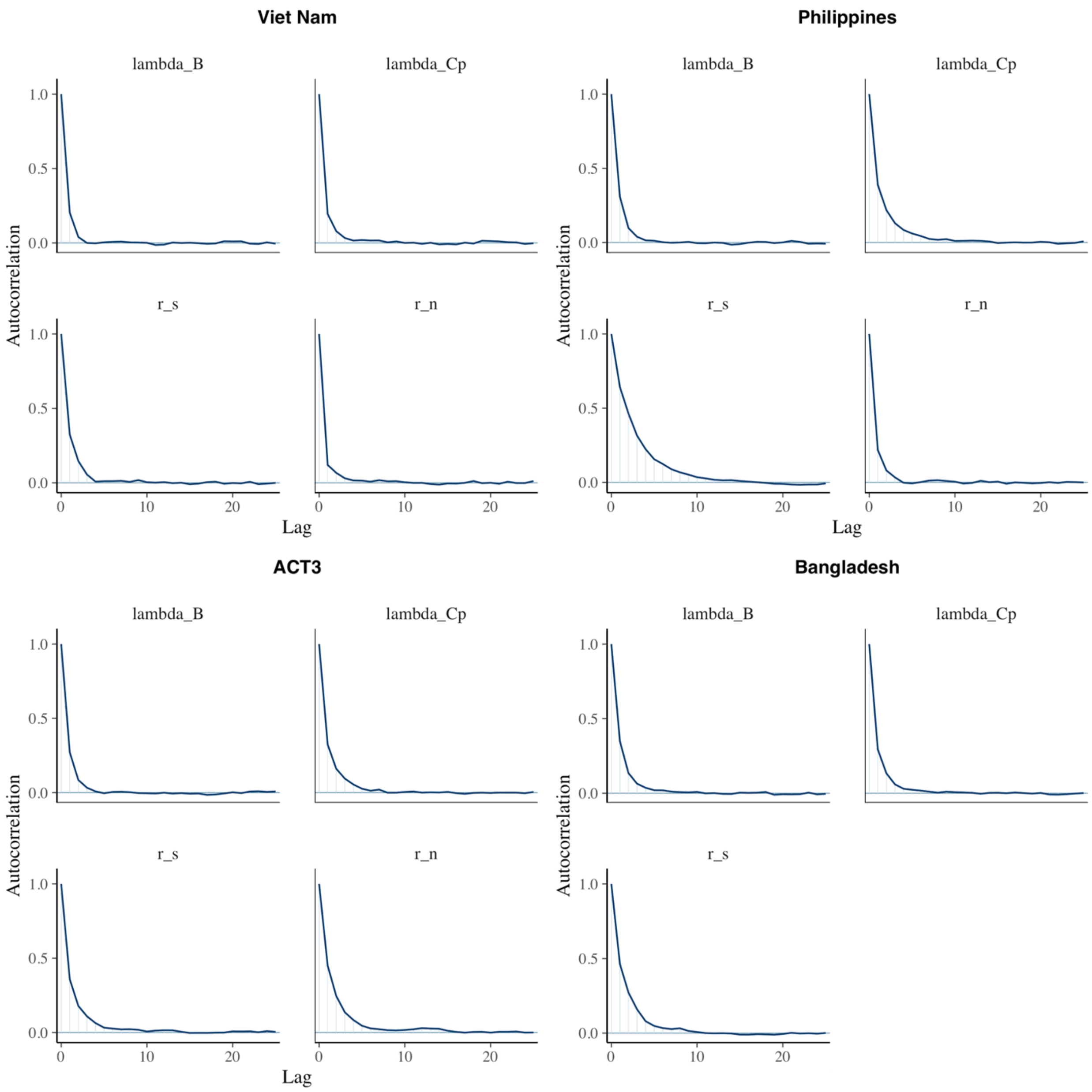


**Supplementary Figure 5:** Autocorrelation plots for each model.

#### *Sensitivity analyses*

*Sensitivity analysis 1:* Omitting Bangladesh (2007) [3] and ACT3 (2017) [5] from the analysis.

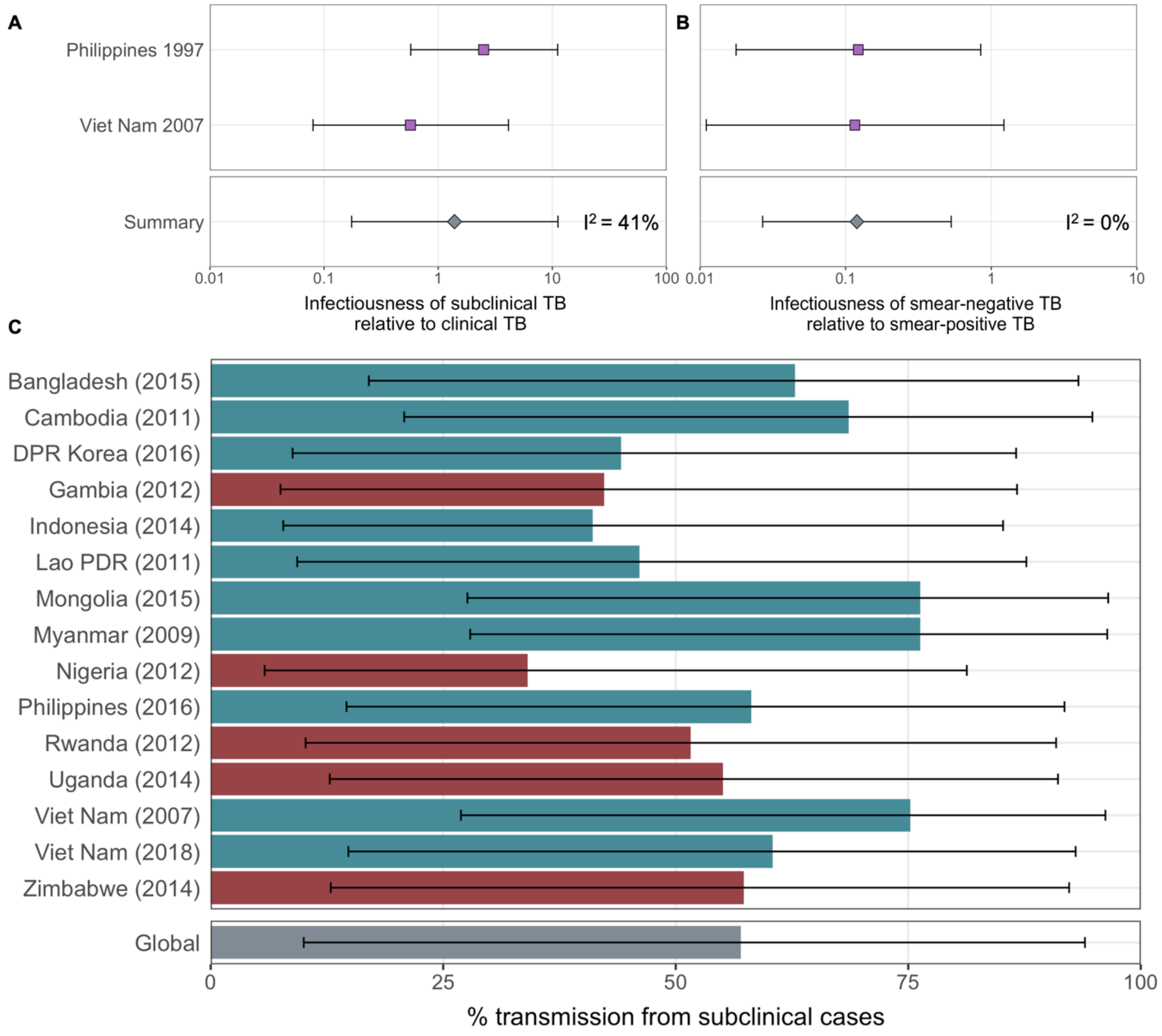


**Supplementary Figure 6:** Affected results for Sensitivity Analysis 2. Figure details are as per **Figure 2A-B** and **Figure 3D** in the main text.

*Sensitivity analysis 2:* Using an alternative duration of subclinical TB relative to clinical TB from [22].

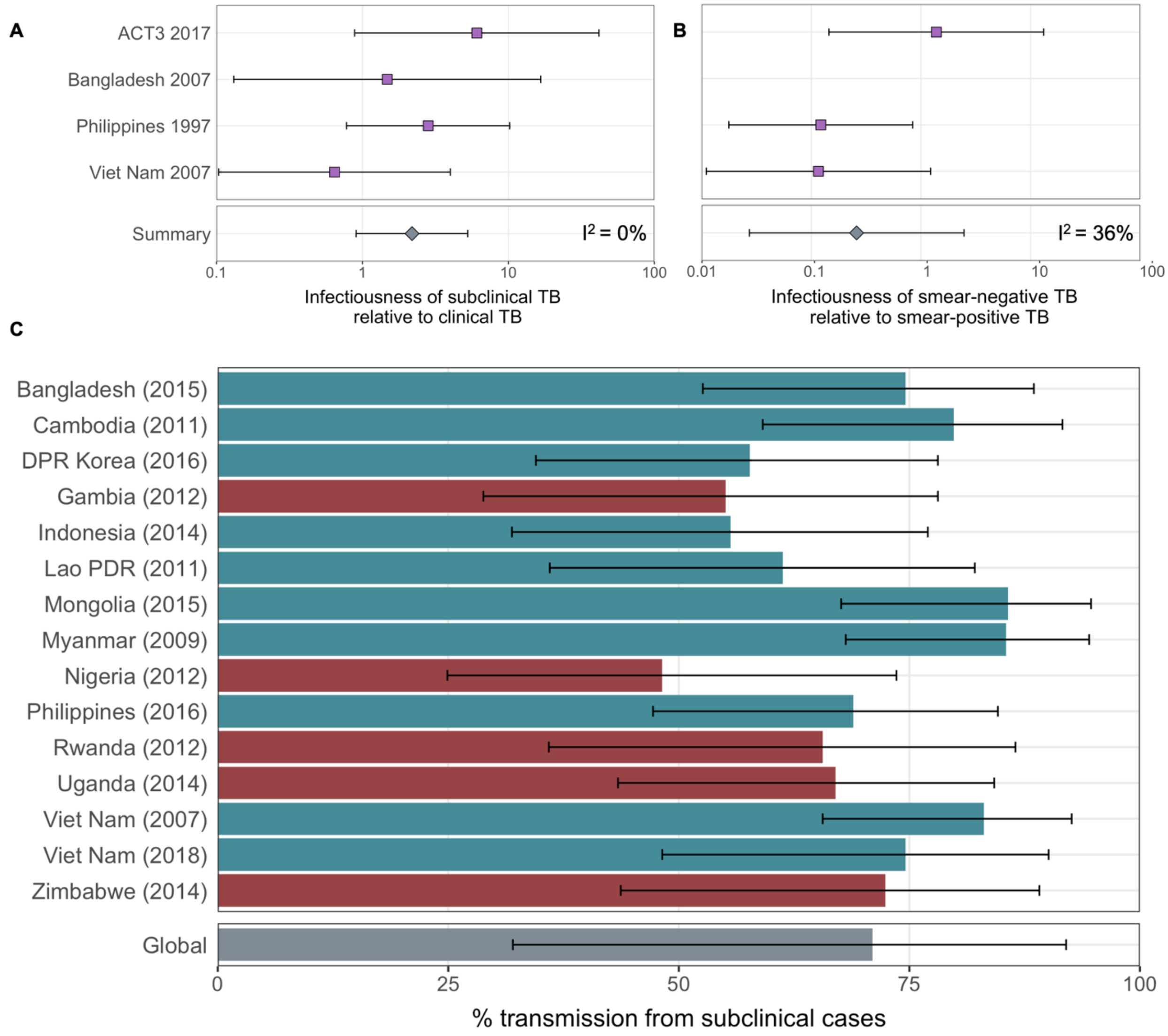


**Supplementary Figure 7:** Affected results for Sensitivity Analysis 2. Figure details are as per **Figure 2A-B** and **Figure 3D** in the main text.

11 Ministry of Health – Cambodia. Report of the Second National Tuberculosis Prevalence Survey, 2011. 2012.

12 Ministry of Health and Social Welfare - The Gambia. First Ethiopian National Population Based Tuberculosis Prevalence Survey. 2011.

13 Ministry of Health – Rwanda. Report of the First National Pulmonary Tuberculosis Prevalence Survey in Rwanda. 2014.

14 Federal Republic of Nigeria. Report: First National TB Prevalence Survey. 2012.

15 Ministry of Health, Republic of Indonesia, National Institute of Health Research And Development In collaboration with Directorate General of Disease Control and Environmental Health. Indonesia Tuberculosis Prevalence Survey 2013-2014. 2015.

16 The Republic of Uganda. The Uganda National Tuberculosis Prevalence Survey, 2014-2015 Survey Report.

17 Ministry of Health and Child Care – Zimbabwe. Report: The Zimbabwe National Population Based Tuberculosis Prevalence Survey. 2014.

18 DGHS Ministry of Health and Family Welfare - Bangladesh. National Tuberculosis Prevalence Survey Bangladesh 2015-2016.

19 Ministry of Health – Mongolia. Report of the First National Tuberculosis Prevalence Survey in Mongolia 2014-2015. 2016.

20 Democratic People's Republic of Korea. National TB Prevalence Survey 2016.

21 Department of Health - Philippines. National Tuberculosis Prevalence Survey 2016.

22 Richards AS, Sossen B, Emery JC, *et al.* The natural history of TB disease-a synthesis of data to quantify progression and regression across the spectrum. 2021. doi:10.1101/2021.09.13.21263499
